# USING MACHINE LEARNING OR DEEP LEARNING MODELS IN A HOSPITAL SETTING TO DETECT INAPPROPRIATE PRESCRIPTIONS: A SYSTEMATIC REVIEW

**DOI:** 10.1101/2023.06.01.23290818

**Authors:** E. Johns, J. Godet, A. Alkanj, M. Beck, L. Dal Mas, B. Gourieux, E.-A. Sauleau, B. Michel

## Abstract

**Objectives:** The emergence of artificial intelligence (AI) is catching the interest of hospitals pharmacists. Massive collection of pharmaceutical data is now available to train AI models and hold the promise of disrupting codes and practices. The objective of this systematic review was to examine the state of the art of machine learning or deep learning models that detect inappropriate hospital medication orders.

**Methods:** A systematic review was conducted according to the PRISMA statement. PubMed and Cochrane database were searched from inception to May 2023. Studies were included if they reported and described an AI model intended for use by clinical pharmacists in hospitals.

**Results:** After reviewing, thirteen articles were selected. Eleven studies were published between 2020 and 2023; eight were conducted in North America and Asia. Six analyzed orders and detected inappropriate prescriptions according to patient profiles and medication orders, seven detected specific inappropriate prescriptions. Various AI models were used, mainly supervised learning techniques.

**Conclusions:** This systematic review points out that, to date, few original research studies report AI tools based on machine or deep learning in the field of hospital clinical pharmacy. However, these original articles, while preliminary, highlighted the potential value of integrating AI into clinical hospital pharmacy practice.

**What is already known on this topic:** AI models for pharmacists are at their beginning. Pharmacists need to stay up-to-date and show interest in developing such tools.

**What this study adds:** This systematic review confirms the growing interest of AI in hospital setting. It highlights the challenges faced, and suggests that AI models have a great potential and will help hospital clinical pharmacists in the near future to better manage review of medication orders.

**How this study might affect research, practice or policy:** AI models have a gaining interested among hospital clinical pharmacists. This systematic review contributes to understand AI models and the techniques behind the tools.

## Introduction

Clinical pharmacy is a health science discipline in which pharmacists provide patient care that optimizes medication therapy and promotes health, and disease prevention (1). Computerization of the medication use process (from prescribing to administration) allowed clinical pharmacists to access prescriptions more easily and to perform tasks that make medication use safer, including the review of medication orders. With this computerization, tools were then developed such as computerized clinical decision support systems (CDSS) dedicated to drug alerts. CDSSs are intended to improve patient safety by assisting clinicians in making decisions. It can help pharmacists in their task, but despite the benefits, it also has pitfalls. CDSS fragments the workflow of the user, but also over alerts for non-relevant or inappropriate signals leading to fatigue (desensitization) and subsequent inefficiency (2). A drug alert CDSS is usually interfaced with a national drug database. These systems alert the prescriber and/or the pharmacist if a prescription is inappropriate, based on the implemented rules. These are usually based on the summaries of product characteristics and/or other validated drug databases available.

The computerization of health data has led to a large-scale collection of data. This massive quantity of data has become a powerful mine for the development of new tools to help healthcare professionals in their clinical practice. Technologies using artificial intelligence (AI) need massive collection of data to be generated. Such applications are already developed and in use in imaging (3) or cancer prediction and diagnosis (4) for examples.

AI is the ability of a machine to display human-like capabilities such as learning and classifying. AI is a vast field regrouping several techniques, such as machine learning (ML). ML systems are designed to define its own set of rules based on data during training. The main tasks of ML are classification, regression, clustering, dimension reduction, or association. By defining their own rules, these algorithms will predict the probability of the outcome occurring on new data, generalizing from previously learnt data. The different models are based on the available data and the aim of the prediction. The learning methods are multiple: supervised (based on a labeled dataset indicating the expected outcome), unsupervised (based on the selection of relevant features between data of a given dataset), semi-supervised (based on mixing labeled and unlabeled data in the training set), self-supervised (based on learning the input data from another part of the input dataset) or also reinforced (based on interacting with the environment: reinforcement by a reward/punishment system) (5). Deep learning (DL) is a subset of ML, based on a neural network composed of hidden layers selecting distinguishing features of the training dataset (6). DL mimics the complexity of the human decision process.

These AI technologies definitely offer the potential to disrupt clinical practices by developing tools to help pharmacists prioritize medication order reviews for high-risk patients, to facilitate decision-making process in drug selection, to predict dosage of narrow therapeutic index drugs, drug-drug interactions or adverse drug reactions, to enable efficient pharmacy workforce allocation in a resource-constrained environment.

In light of such prospects, there is currently a growing interest among research teams to develop AI tools to assist clinical pharmacists in their daily practice. Two literature reviews (7,8) have gathered information on such studies and have highlighted the potential value of integrating AI into pharmacy practice. Our study complements these reviews by adding the latest developed models and focusing specifically on AI-derived tools designed to detect inappropriate prescribing in the hospital setting for clinical pharmacists.

## Objective

The goal of this systematic review was to summarize the existing literature on predictive algorithms to detect inappropriate medication orders, in a hospital setting and using ML or DL technologies.

## Methods

### Eligibility criteria

To enter the systematic review, the inclusion criteria were the development and description of a ML or DL algorithm detecting inappropriate prescription in a hospital setting. Articles had to be written either in English or in French and be peer-reviewed and published in a journal.

### Information sources

The review was conducted according to the Preferred Reporting Items for Systematic Reviews and Meta-Analysis (PRISMA) statement (9). PubMed database and the Cochrane Library were searched from inception to May 2023.

### Search strategy

A targeted search of the literature was done, to analyze the vocabulary and choose the appropriate keywords for our review. The following terms (MeSH terms associated with complementary terms) were selected:

- “artificial intelligence” AND “clinical decision support system”,
- “artificial intelligence” AND “clinical pharmacy”,
- “clinical pharmacy” AND “clinical pharmacy information systems”,
- “computerized” AND “clinical decision support systems”,
- “computerized” AND “pharmacy data”,
- “machine learning” AND “clinical decision support system”,
- “pharmaceutical” AND “algorithm”,
- “pharmaceutical” AND “decision support system”,
- “pharmacy” AND “machine learning”,
- “pharmacy” AND “deep learning”.

### Selection process

To enter the selection process, the articles were screened based on their title. To be selected, they had to address the use of AI for clinical pharmacists in the hospital setting. After this initial selection, the articles were screened based on their abstract, which had to detail the AI technology used. Finally, the selection was based on full texts. Were excluded articles reviews, articles not oriented towards the detection of inappropriate prescriptions, not hospital-based articles and non-available articles (access or language issues).

### Data collection process

Extracted data included: references of the article, objectives, description of the AI model, dataset used, main results, contributions and limitations of the model.

Selection and data extraction were conducted by one reviewer.

### Study risk of bias assessment

This systematic review has a risk of bias since the first selection process was done based on the title of the article. Articles without an explicit title were not included in the study.

## Results

### Identification and selection of studies

The queries identified 504 articles. 316 articles, after abstract screening, did not meet the inclusion criteria. Of the 188 remaining articles, 175 were excluded for various reasons: review articles (n=20); not relevant (n=101) because no AI was used or the model used was not described but only evaluated; not oriented towards inappropriate orders detection (n=35) but used for pharmacovigilance surveillance; not hospital-based (n=8); not accessible (n=6); or written in a language other than English or French (n=5). Finally, 13 articles were included in the systematic review. (Figure 1)

**Figure 1:**
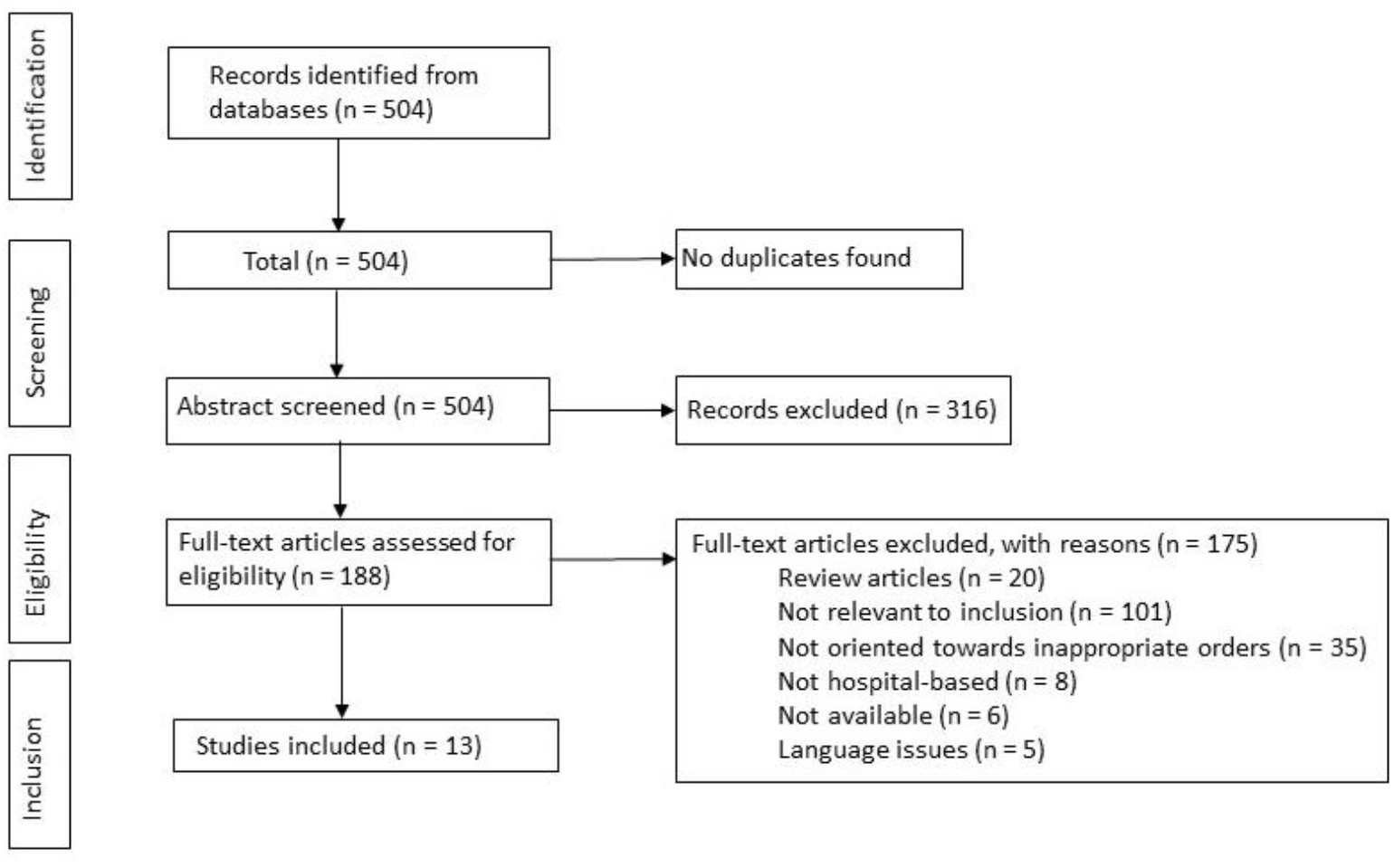
PRISMA flow diagram of the selection of the included studies

### General description of the articles

The studies were mainly conducted in North America (n= 4) and Asia (n=4). Eleven articles (84.6%) were published between 2020 and 2023.

Six articles aimed to analyze orders and detect inappropriate prescriptions according to patient profiles and medication orders:

- Prediction of high risk of QT prolongation due to drug interactions (10),
- Prediction of orders requiring an intervention after analyzing the order provider’s interaction with the electronic health record (11),
- Characterization of the risk factors associated with medication ordering errors (12),
- Detection of medication errors in neonatal intensive care unit (13),
- Identification of atypical medication orders and pharmacological profiles (14),
- Prioritizing of medication order reviews to reduce the risk of errors (15).
- Two articles (16,17) were oriented towards antibiotic resistance detection:
- Detection of antimicrobial inappropriate prescriptions (16),
- Prediction of antibiotic resistance on bacterial infections for five different antibiotics (17). Two articles (18,19) presented algorithms predicting the risk of adverse drug events (ADE):
- Prediction of ADE for elderly patients (18),
- Prediction of the risk of ADE for individual patients (19).
- Two articles (20,21) described an algorithm detecting dosage abnormality in prescriptions:
- Detection of prescription outliers (wrong dosage and frequency) (20),
- Detection of extreme overdosing or underdosing in prescriptions (21).

Finally, one article (22) described an algorithm capable of screening high alert drugs errors from prescriptions.

### Algorithmic models description

Eight publications (10–13,17–19,22) used supervised ML models (Table 1) and unsupervised ML algorithms (Table 2) were used in three publications (14,20,21).

**Table 1:** Algorithm using supervised learning models

| Reference | Outcome | AI model | Dataset | Main results |  |  |  |  |  |  |  | Contributions | Limitations |
| --- | --- | --- | --- | --- | --- | --- | --- | --- | --- | --- | --- | --- | --- |
|  |  |  |  | Characteristics | Acc | Rec | Prec | Spe | F1 | AUROC | AUCPR |  |  |
| Machine Learning Techniques Outperform Conventional Statistical Methods in the Prediction of High Risk QTc Prolongation Related to a Drug-Drug Interaction (Van Laere S., et al., Belgium, 2022) (10) | Develop an algorithm predicting high risk of QTc prolongation and alert when DDIs increase the risk of QTc prolongation | Linear regression, LR, Gaussian Naïve Bayes classifier, DT, SVM, RF, boosting algorithms | **Training set:** 512 QT-DDIs (284 low risk QT-DDIs, 228 high risk QT-DDIs, 289 male patients, 223 female patients).<br>**Hold-out set:** 102 QT-DDIs (57 low risk QT-DDIs, 45 high risk QT-DDIs, 52 male patients, 50 female patients) | RF | 0.82 | 0.76 | 0.83 | 0.88 | / | / | / | ML techniques have higher performances as conventional statistical methods, especially random forest and Adaboost models. The analyze of the top-8 features used in the classification helps explain the model. The training set was stratified with 5 different folds, balancing the predictive model used in the validation set. No | The small dataset used and requiring a strict QTc monitoring. The validation set was predicted from the strongest performing model. Missing values were imputed with the median of the known values. |
|  |  |  |  | Adaboost | 0.82 | 0.73 | 0.83 | 0.88 |  |  |  | indication of the DDI was modeled making the algorithm generalizable to another sample of QT-DDIs. |  |
| Predicting inpatient pharmacy order interventions using provider action data (Balestra M., et al., USA, 2021) (11) | Predict orders requiring intervention from the ordering provider's interaction with the EHR | XGBoost | 1811407 individual orders, from 2708 prescribers, extracted from 3 hospitals |  | 0.41 | 0.99 | / | 0.37 | / | 0.91 | 0.44 | With proper tuning, such models can significantly improve the workloads on pharmacists. | Dataset extract from a short period (2 weeks), seasonality of behaviors may be explored. Comparison of the results across hospital systems to explore the context influence on behavior. |
| Predicting self-intercepted medication ordering errors using machine learning (King C. R., et al, USA, 2021) (12) | Characterize the risk factors associated with medication ordering errors | LR-DT-RF-MLP-GBDT | Medication orders (+associated data) over a 6-year period in a single hospital (58041920 orders - 28695 voided orders) | GBDT |  |  |  |  |  | 0.80 | 0.07 | Large collection of data. GBDT model has the best performance. Identification of factors associated with order errors and of medication orders in a high-risk context. | The performance of the models is below the standards expected to use such models in clinical practice. Generalizability to other data not studied. |
| Development and validation of a machine learning-based detection system to improve precision screening for medication errors in the neonatal intensive care unit (Yalçın N. et al., Türkiye, 2023) (13) | Develop a ML model predicting the presence of medication errors | RF | Prospective study, included 412 NICU-patients to whom at least one systemic drug was prescribed over a 17-month period in a single hospital (11 908 medication orders) |  | 0.92 | 0.92 | 0.94 | 0.92 | 0.93 | 0.92 | / | First developed and validated model to predict the presence of medication errors using work environment and pharmacotherapy parameters. Expectation to predict the occurrence of a medication error without causing alert fatigue. RF had the highest performance. | Small and single unit hospital dataset limiting the heterogeneity of the data pool and the generalizability to other populations. |
| Predicting Antibiotic Resistance in Hospitalized Patients by Applying Machine Learning to Electronic Medical Records (Lewin-Epstein O., et al., Israel, 2021) (17) | Predict the antibiotic resistance of bacterial infections to the five antibiotics tested for resistance: ceftazidime, gentamicin, imipenem, ofloxacin, sultrim | Ensemble-based ML (LR, GBDT, Neural network) | Electronic medical records of patients hospitalized in a single hospital, over a 32-month period: 16000 antibiotic-resistance tests of bacterial cultures | Identity of bacterial species known | 0.70 | / | / | / | / | 0.76 | / | Ability to predict antibiotic resistance from a large relatively incomplete dataset. An ensemble of three algorithms produces robust results, without the pitfalls of single algorithms. | Additional information in the EMR could improve the patient's profile thus the resistance prediction. The model should be periodically retrained to reflect the resistance patterns and antibiotic consumptions. |
|  |  |  |  | Identity of bacterial species unknown | 0.74 | / | / | / | / | 0.82 | / |  |  |
| Predicting adverse drug events in older inpatients: a machine learning study (Hu Q. et al., China, 2022) (18) | Develop a model to predict ADE in older inpatients | XGBoost, AdaBoost, CatBoost, GBDT, LightGBM, TPOT, RF | 1880 randomly selected patients over a year period | Adaboost | 0.88 | 0.43 | 0.69 | / | 0.53 | / | / | ML techniques catch the complex relationships between the variables and learn from data situations. Adaboost model has high performance results. | Small and single-centered sample. The retrospective review of the medical records depends on the documentation quality. Adding more information to the algorithm could help predict ADE more accurately. |
| Validation of the usefulness of artificial neural networks for risk prediction of adverse drug reactions used for individual patients in clinical practice (Imai S., et al., Japan, 2020) (19) | Validate the usefulness of ANNs using MLP algorithm to predict the risk of ADRs for an individual patient | ANNs: MLP | 1141 subjects who had received Vancomycin intravenously, over a 7-year period in a single hospital |  | 0.86 | / | / | / | / | 0.83 | / | MLP have slightly better results than logistic regression, thus enabling better prediction. ANN models would help clinicians in drug selection and avoiding ADRs. | Small dataset (single center study). Factors included in study without evaluation at the time. |
| High alert drugs screening using gradient boosting classifier (Wongyikul P., et al., Thailand, 2021) (22) | Develop a ML model and a HAD screening protocol aiming the appropriateness of HAD use: screening HAD errors from drug prescriptions | Gradient booster classifier | OPD: 991270 prescriptions, 3280 unique ICD10 codes, 1184 unique drug codes; IPD: 1200000 prescriptions, 2020 unique ICD10 codes, 1767 unique drug codes | HAD binary classification: OPD dataset | 0.75 | 0.83 | 0.23 | / | / | 0.36 | / | Potential benefit to reduce the manual drug verification process and precisely verify the appropriateness between high alert drug prescriptions and ICD10s. | Manual final check process of HAD-ICD10 mismatches and relies on a clinical judgment. Insufficient number of some HADs and ICD10s. Not able to scale to a large amount of data. Only use of information between drugs and ICD10s to evaluate the appropriateness of HAD use. |
|  |  |  |  | HAD binary classification: IPD dataset | 0.69 | 1.00 | 0.67 | / | / | 0.80 | / |  |  |
|  |  |  |  | HAD type classification: OPD dataset | 0.64 | 0.93 | 0.12 | / | / | 0.20 | / |  |  |
|  |  |  |  | HAD type<br>classification:<br>IPD dataset | 0.59 | 0.93 | 0.20 | / | / | 0.32 | / |  |  |

Acc: Accuracy; ADE: Adverse Drug Event; ADR: Adverse Drug Reaction; ANN: Artificial Neural Network; AUCPR: Area Under the Precision-recall Curve; AUROC: Area Under the Receiver Operating Characteristic; DDI: drug-drug interaction; DT: Decision Tree; EHR: Electronic Health Record; EMR: Electronic Medical Records; F1: F1-score; GBDT: Gradient Boosted Decision Tree; HAD: High Alert Drug; ICD10: International Classification of Diseases 10th Revision; IPD: InPatient Department; LightGBM: Light Gradient-Boosting Machine; LR: Logistic Regression; ML: Machine Learning; MLP: MultiLayer Perceptron; NICU: Neonatal Intensive Care Unit; OPD: OutPatient Department; Prec: Precision; QTc: corrected QT interval; Rec: Recall; RF: Random Forest; Spe: Specificity; SVM: Support Vector Machine; TPOT: Tree-based Pipeline Optimization Tool;

**Table 2:** Algorithm using unsupervised learning models

| Reference | Outcome | AI model | Dataset | Main results |  |  |  |  |  |  |  | Contributions | Limitations |
| --- | --- | --- | --- | --- | --- | --- | --- | --- | --- | --- | --- | --- | --- |
|  |  |  |  | Characteristics | Acc | Rec | Prec | Spe | F1 | AUROC | AUCPR |  |  |
| Pharmacists' perceptions of a machine learning model for the identification of atypical medication orders (Hogue S.-C., et al., Canada, 2021) (14) | Identify atypical medication orders and pharmacological profiles | GANomaly-based model (self-learner) | 12624 medication orders and 2114 pharmacological profile, over a 4-month period | Medications orders | / | 0.26 | 0.35 | 0.97 | 0.30 | 0.80 | 0.25 | Better performance of the model to identify atypical pharmacological profiles than atypical medication orders. Pharmacists seemed to find medication order prescriptions more useful. | Medication ordering patterns may include suboptimal but common practices. This model should rather be combined with classical rule-based approaches to detect such issues independently of practice patterns. Single-center study. |
|  |  |  |  | Pharmacological profiles | / | 0.75 | 0.49 | 0.82 | 0.59 | 0.88 | 0.60 |  |  |
| DDC-Outlier: Preventing Medication Errors Using Unsupervised Learning (Santos H., et al., Brazil, 2019) (20) | Detect automatically wrong dosages and frequencies for medications in electronic prescriptions: detection of prescription outliers | Density distance centrality algorithm (Jaccard similarity) | 240000 orders, 2 million medications, 16000 patients, over a 9-month period | / | / | 0.90 | 0.61 | / | 0.68 | / | / | The algorithm could detect other prescriptions to be improved by the pharmacy department. The outlier detection of prescriptions with homogeneous prescription distribution was better, than sparse prescription distribution. | Medicine depending on the patient's weight were discarded to avoid a false outlier, because the patients' weights were not available in the dataset. The algorithm detects overdose/ |
|  |  |  |  |  |  |  |  |  |  |  |  |  | underdose as the unique medication problem. No detection of non-standard prescriptions. |
| Detection of overdose and underdose prescriptions- An unsupervised machine learning approach (Nagata K., et al., Japan, 2021) (21) | Detect prescription errors of overdoses and underdoses using ML: detect extreme overdose and underdose prescriptions that occur rarely in clinical practice | One-class SVM | 31 clinical overdose and underdose prescriptions of oral drugs (21 drugs, > 1000 in-hospital prescriptions) were analyzed. 87% were detected as abnormal. | Overdose prescriptions | / | 0.96 | 0.99 | / | 0.97 | / | / | "Age" and "weight" parameters are crucial when detecting dose prescription errors. The addition of synthetic data increased the performance of the model. | Insufficient clinical overdoses and underdoses data were used to evaluate the model's performance. The model may not be able to detect rare prescription errors. The |
|  |  |  |  | Underdose prescriptions | / | 0.79 | 0.98 | / | 0.84 | / | / |  | detection of underdose prescription errors were slightly lower than the overdose prescription errors. Additional factors affecting dosage should be added. |

Acc: Accuracy; AUCPR: Area Under the Precision-recall Curve; AUROC: Area Under the Receiver Operating Characteristic; DDC: Density-distance-Centrality; F1: F1-score; ML: Machine learning; Prec: Precision; Rec: Recall; Spe: Specificity; SVM: Support Vector Machine

To optimize the performance of a rule-based system, combining ML models with such systems (Table 3) was explored in two publications (15,16).

**Table 3:** Algorithm using hybrid models

| Reference | Outcome | AI model | Dataset | Main results |  |  |  |  |  |  |  | Contributions | Limitations |
| --- | --- | --- | --- | --- | --- | --- | --- | --- | --- | --- | --- | --- | --- |
|  |  |  |  | Characteristics | Acc | Rec | Prec | Spe | F1 | AUROC | AUCPR |  |  |
| A machine learning-based clinical decision support system to identify prescriptions with a high risk of medication error (Corny J., et al., France, 2020) (15) | Prioritize prescription reviewing to reduce the risk of prescribing errors | LightGBM, a gradient-boosting framework based on DT algorithms + rule-based expert system | DxCare, national drug database, Thériaque, published literature, 133179 prescription orders (medication order data, laboratory reports, demographics, medical history, vital signs), over a 18-month period | / | / | 0.74 | 0.74 | / | 0.74 | 0.71 | 0.75 | Prediction at the patient level. Algorithm outperforms classic systems in detecting medication errors. The use of a hybrid system could help identifying critical medical errors (never-events) and reduces the number of false alerts. | Study conducted in a single hospital, excluding neonatology and intensive care unit patients. More pharmaceutical interventions were recommended during the test phase than in the development phase. No real-life evaluation. |
| An Antimicrobial Prescription Surveillance System that Learns from Experience (Beaudoin M. et al., Canada, 2014) (16) |  | Hybrid model: rule induction + kNN | 5756 patients, 19172 antimicrobial prescriptions (7027 prescriptions triggered an alert), over a 6-month period | TAZO dataset | 0.74 | 0.99 | 0.63 | / | / | / | / | Specific-to-general search and modification of every rule in parallel. Clinical relevance of learned rules. | Imbalanced dataset may complicate the learning of inappropriate prescriptions. |
|  |  |  |  | METRO dataset | 0.85 | 0.76 | 0.54 | / | / | / | / |  |  |

Acc: Accuracy; AUCPR: Area Under the Precision-recall Curve; AUROC: Area Under the Receiver Operating Characteristic; DT: Decision Tree; F1: F1-score; kNN: k-Nearest Neighbor; LightGBM: Light Gradient-Boosting Machine; METRO: METRONidazole; ML: Machine Learning; Prec: Precision; Rec: Recall; Spe: Specificity; TAZO: piperacillin-TAZObactam;

These models were evaluated using different metrics. Commonly the following metrics used are (23,24):

- Accuracy: ratio between correctly classified samples and the total samples of the evaluation set,
- Recall: also called sensitivity or true positive rate, ratio between the true positive samples and the total of positive samples,
- Precision: also called positive predictive value, ratio between correctly classified samples and the total of samples assigned to that class,
- Specificity: also called true negative rate, ratio between true negative samples and the total of negative samples,
- F1-score: harmonic mean of precision and recall,
- Area Under the Receiver Operating Characteristic (AUROC): measure of the overall performance of the test,
- Area Under the Precision-Recall Curve (AUCPR): summary of the precision-recall curve.

### Supervised ML models

Eight articles (10–13,17–19,22) described the use of supervised ML. Five ML models were tested: DT (10,12,13,18), SVM (10), bagging (10,12,18), boosting (10–12,17,18,22) and neural network (12,17,19). Three articles (11,19,22) compared distinct ML techniques. Lewin-Epstein et al (17) combined multiple techniques to enhance the results. Three articles (10,12,22) compared ML algorithms with conventional statistical methods.

### Unsupervised ML models

Three articles (14,20,21) reported the use of unsupervised ML. Each study described a specific model: Jaccard similarity, Support Vector Machine and GANomaly-based model. The datasets contained less prescription orders than in the studies using supervised ML. The same metrics were used to evaluate the model (recall, precision, F1-score), but Hogue et al reported additional results (specificity, AUROC, AUPR). The results were heterogeneous: Santos et al and Nagata et al had similar performances (F1-score of 0.68 and 0.97 respectively), whereas Hogue et al had lower results (F1-score of 0.30 for the identification of atypical medication orders and an F1-score of 0.59 for identifying atypical pharmacological profiles).

### Hybrid models

Two selected articles (15,16) presented hybrid models, meaning the algorithm mixed ML techniques with selected rule-based data. Both studies used supervised ML algorithms: boosting and k-nearest-neighbor classification methods.

## Discussion

This literature review identified research articles presenting algorithms developed in a hospital setting for inappropriate medication orders detection. It is still too early to predict the impact of such AI-tools in real clinical pharmacy practice conditions, but this review underlines the importance of addressing this issue and highlights the multiplicity of prediction methodological approaches used.

The diversity of the models developed attests the many possibilities of AI. Supervised ML were meanly used because they are well adapted to classification problems. Boosting and bagging methods, seem to have the best results (10,12,13,18). The lack of results-metric standardization makes it hard to compare the studies (25). It is indeed challenging to confront algorithms across studies because different measures were reported to summarize the AI-tools performance and the metrics were not based on the same datasets and contexts. For example, some studies only published “AUCPR-AUROC” results (12), whereas other teams used “recall-precision-F1-score” (10,16,18,20–22) or both metrics (11,13–15). In addition, training models on a defined dataset questions the generalization and applicability of the model on a new dataset. Most of the studies were single-centered, meaning the data used were representative of only one specific hospital. Implementing models trained on a specific dataset in a new setting, may require model adjustments before using the model routinely. Furthermore, the use of retrospective data, including incorrect, non-relevant or partial data, increases the risk of reproducing these errors. Processing the data before training is challenging and time consuming, but essential to reflect and adapt the set to reality and validate the extraction. The quality of the data determines the performance of the model (26–28). Hybrid models can help avoid these limitations by adding rules to counter the lack of data and its quality in the dataset.

Today, only the hybrid models of this review are used in daily practice (15,16), probably due to their facilitated acceptance by the clinical pharmacists since some predicting parameters are more controllable. However, such rule-based systems imply a continuous manual-update of the base to reflect up-to-date knowledge, whereas the predictive models could update themselves thanks to reinforcement learning methods (29). Clinical pharmacists should be able to understand how the proposed algorithms can improve patient care within a realistic workflow, but most articles do not attempt to present such information. Guidelines and recommendations are emerging, defining a global framework to regulate and provide advice to research and development teams (30,31). Also, Lundberg et al developed a method to explain the output of a model: the SHAP (Shapley Additive exPlanations) value, by averaging the importance of each variable on the model for each possible combinations of variables (32). Explainability and auditability of the technical algorithms will help the clinical pharmacists gain confidence and understanding of the differences in the models (33).

Also, pharmacists should integrate computerization to their trainings in order to understand and acquire knowledge in clinical informatics to use such tools and be aware of the limits and the absolute need of an expert validation of the outcome (34). These initiatives for training programs in health informatics and AI fundamentals, described in the study of Tsopra et al, have been highly valued by undergraduate medical students but also by senior clinicians (35). Developers must also grasp the issues faced and tasks completed by the pharmacists: defining the requirements, the choice of the model and the dataset are the basis for developing functional AI-models. Without interdisciplinary teams, the development of AI models will be compromised. However, pharmacists must be aware that the tool is not perfect and a critical mind and pharmaceutical knowledge are still necessary to validate the prediction, as false positives and false negatives results will still occur.

Deploying fully AI tools is challenging. The models must be registered as a medical device and therefore meet specific criteria, including risk analysis or clinical evaluation. Beyond demonstrating the benefit of the tool, the use of AI complicates the marketing authorization. The competent authorities are legislating on the implementation and use of these tools (36,37), to ensure a safe, high quality and trustworthy AI in a lifecycle regulatory framework. From the design of the model to its marketing, analysis of the quality of the data, listing of potential bias, technical robustness and human supervision are essential. Accountability, regulation and ethical approaches will be shared between model developers and end users.

A major limit of our study is that commercial solutions were not included in this review because of the lack of research and development methods publicly available. Research in AI is mainly carried out by private companies, thus limiting the number of publications. However, commercial solutions are evaluated by clinicians (38) and prove to benefit the patients’ care. The lack of explication of solutions is understandable and ensures the preservation of property rights of the technology. Nonetheless, this contributes to maintaining the opacity around AI for pharmacists and their reluctance to use such tools (39). Shifting this research field to independent and academic teams will reduce the pharmacists’ hesitation to use these models and help ensure wide availability and easy access to the results and characteristics of the algorithms. Open science disseminates good practice and knowledge about the advantages but also limits of AI, giving pharmacists the tools to take a critical look at these commercial solutions and make informed choices about how to integrate them into their practices.

The profession of clinical hospital pharmacists is moving towards a practice assisted by AI, to improve clinical practices for more safety, to optimize the effectiveness of pharmaceutical human resources and the medico-economic efficiency of hospitals. As described in our review, AI will be becoming more and more a technological companion for pharmacists and will bring help to these professionals. Indeed, the data analysis capabilities far exceed human possibilities, allowing much more advanced levels of analysis. Nevertheless, beware, the pharmacist must and will always remain the decision-maker. This is the limit of this new technology, which must help more than decide. By analyzing large amounts of data and synthesizing them for the clinical hospital pharmacists, algorithms must keep their role as companions. In hospitals and in pharmacy in particular, human competence must remain the final step before making a decision. One would have to be oblivious to the changes underway not to imagine tomorrow integrating AI into the daily practice of clinical pharmacy. Technologies are evolving to serve healthcare professionals and patients for greater efficiency and safety.

## Conclusion

The development of AI tools intended for clinical pharmacy practice in a hospital setting is on the rise. The AI models presented have great potential that has to be evaluated and confirmed in further studies and at a large scale. Some clinical pharmacists expressed concerns about consequences on their professional practice and implementation and deployment of AI tools in the hospital setting remain an open question. Hybrid models may be a solution to bridge the uncertainties and guaranty a robust AI-tool. The algorithms presented do not aim at replacing the clinical pharmacist expertise, but provide potential and substantial help to facilitate the organization and workload of the hospital pharmaceutical teams.

## Data Availability

All data produced in the present work are contained in the manuscript.

## Ethical approval

The authors state that no ethical approval was needed.

## Contributorship Statement

Conceptualization: EJ, JG, AA, MB, LDM, BG, EAS, BM

Selection and data extraction: EJ

Data analysis: EJ

Writing of original draft: EJ, JG, BM

Writing review and editing: EJ, JG, AA, MB, LDM, BG, EAS, BM

All authors approved the final version of the work to be published; and agreed to be accountable for all aspects of the work in ensuring that questions related to the accuracy or integrity of any part of the work are appropriately investigated and resolved.

